# Forecasting COVID-19 infection trends in the EU-27 countries, the UK and Switzerland due to SARS-CoV-2 Variant of Concern Omicron

**DOI:** 10.1101/2021.12.16.21267785

**Authors:** Alberto Giovanni Gerli, Stefano Centanni, Joan B Soriano, Julio Ancochea

## Abstract

**Background:** On November 26, 2021, WHO designated the variant B.1.1.529 as a new SARS-CoV-2 variant of concern (VoC), named Omicron, originally identified in South Africa. Several mutations in Omicron indicate that it may have an impact on how it spreads, resistance to vaccination, or the severity of illness it causes.

**Methods:** We used our previous modelling algorithms to forecast the spread of Omicron aggregated in the EU-27 countries, the United Kingdom and Switzerland, and report trends in daily cases with a 7-day moving average. We followed EQUATOR’s TRIPOD guidance for multivariable prediction models. Modelling included a third-degree polynomial curve in existing epidemiological trends on the spread of Omicron in South Africa, a five-parameter logistic (5PL) asymmetrical sigmoidal curve following a parametric growth in Europe, and a new Gaussian curve to estimate a downward trend after a peak.

**Results:** Up to January 15, 2022, we estimated a background rate projection in EU-27 countries, the UK and Switzerland of about 145,000 COVID-19 daily cases without Omicron, which increases up to 440,000 COVID-19 daily cases in the worst scenario of Omicron spread, and 375,000 in the “best” scenario. Therefore, Omicron might represent a relative increase from the background daily rates of COVID-19 infection in Europe of 1.03-fold or 2.03-fold, that is up to a 200% increase.

**Conclusion:** This warning pandemic surge due to Omicron is calling for further reinforcing of COVID-19 universal hygiene interventions (indoor ventilation, social distance, and face masks), and anticipating the need of new lockdowns in Europe.

## Text

Back in March to May 2020, Italy and Spain were the hardest hit European countries during the first wave of COVID-19.^1^ There have been several successful attempts to forecast trends of incidence and mortality of COVID-19, most based upon knowledge on viral dynamics from previous pandemics, recent COVID-19 geographical information of diverse granularity, and newly discovered viral characteristics.^2,3^ However, SARS-CoV-2 inherent poor quality RNAm copy-editing gene replication makes it prone to mutate and spontaneously create new variants of concern (VoC),^4^ that adapt to any hostile environment, produce new outbreaks, and modify existing epidemiological projections.^5^

On November 26, 2021, WHO designated the variant B.1.1.529 as a new VoC, named Omicron, originally identified in South Africa, on the advice of WHO’s Technical Advisory Group on Virus Evolution.^6^ This decision was based on the evidence that Omicron has several mutations that may have an impact on how it spreads, resistance to vaccination, or the severity of illness it causes.^7^ In particular, in South Africa up to December 2, 2021 it was observed a doubling time for the first 3 days after the wave threshold of ten cases per 100 000 population.^8,9^

We used our previous modelling algorithms,^10,11,12^ to forecast the spread of Omicron aggregated in the EU-27 countries, the United Kingdom and Switzerland, and report trends in daily cases with a 7-day moving average. We followed EQUATOR’s TRIPOD guidance for multivariable prediction models.^13^ By applying firstly a third-degree polynomial curve in existing epidemiological trends on the spread of Omicron in South Africa, starting from the first 17 days of the Omicron outbreak (from November 11, 2021), and secondly a five-parameter logistic (5PL) asymmetrical sigmoidal curve following a parametric growth,^10-12^ we were able to model new infections of COVID-19 in South Africa up to December 31, 2021. Overall, the best-case scenario is 80,000 COVID-19 daily cases, while the worst case scenario is 120,000 (Figure 1A).

**Figure 1:**
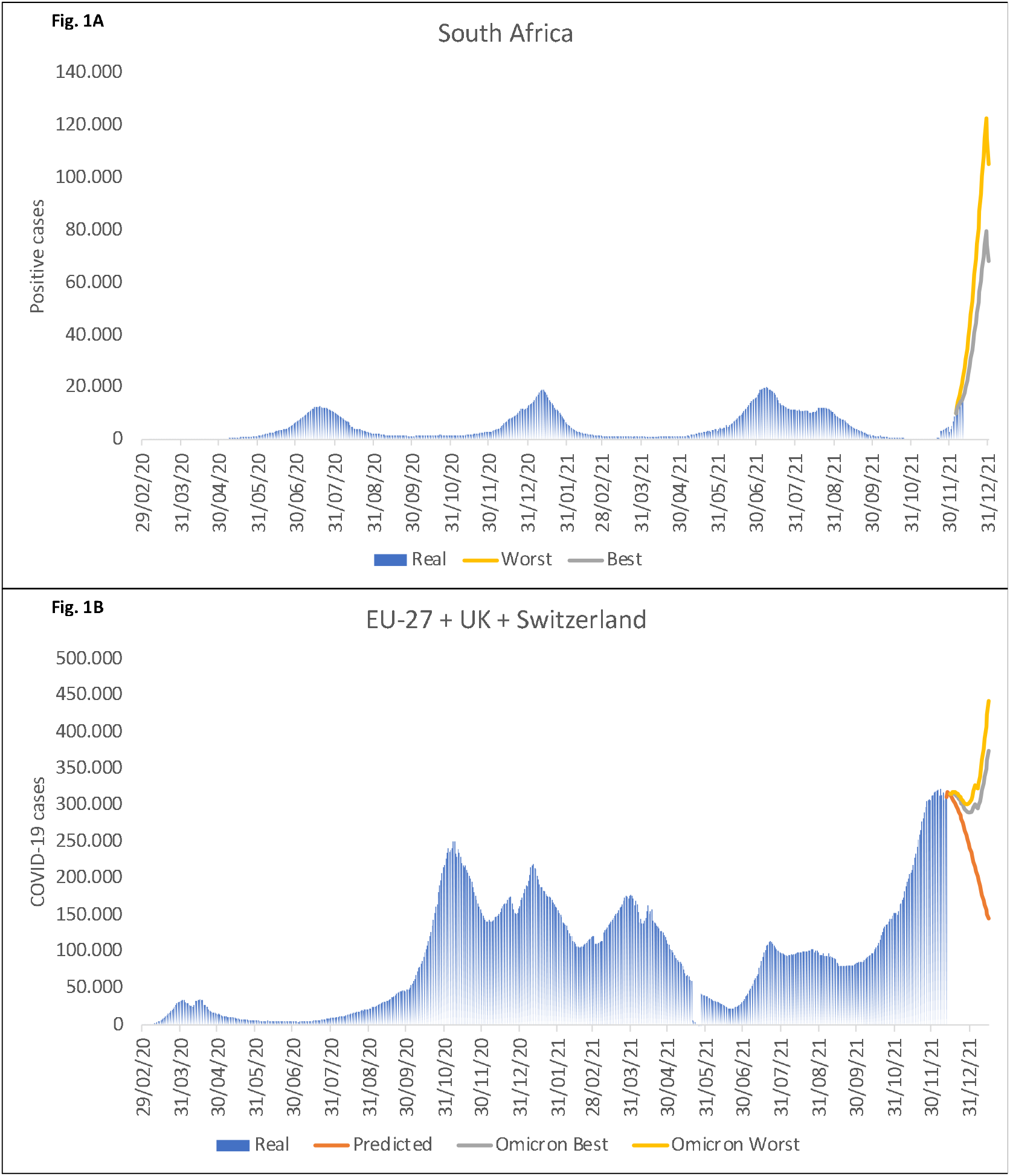
Trends in COVID-19 daily new infections with a seven-day moving average observed and expected in A) South Africa and B) EU-27 countries + UK + Switzerland

Then we modelled these trends to EU-27, the UK and Switzerland new COVID-19 cases using a new Gaussian curve to estimate a downward trend after a peak,^14^ and we obtained the expected curve of new COVID-19 infections in Europe, which does not include the potential effect of Omicron VOC. Finally, we estimated the number of susceptible individuals based on actual vaccination in South Africa and applied them in Europe, and we added those cases to our actual predictive model. Given ongoing trends, we envisage a projection in EU-27 countries, the UK and Switzerland of about 145,000 COVID-19 daily cases by January 15,. 2022 without Omicron, which increases up to 440,000 COVID-19 daily cases by January 15, 2022 in the worst scenario of Omicron spread, and 375,000 in the “best” scenario (Figure 1B). Therefore, Omicron might represent a relative increase from the background daily rates of COVID-19 infection in Europe of 1.03-fold or 2.03-fold, that is up to a 200% increase.

In probability theory, the conditional expectation of any warning system for an eventual surge of an infectious outbreak, as could happen with the Omicron substituting other SAR-CoV-2 VoC, modifies (reduces) the eventual magnitude of the event itself.^15^ Given preliminary evidence from South Africa, our forecast anticipates a large increase in COVID-19 in Europe despite the high levels of vaccination in most of the region. Therefore, this warning is calling for further reinforcing of universal hygiene interventions (indoor ventilation, social distance, and face masks), and anticipating the need of new lockdowns,^11^ the latter being extremely detrimental to many economies.

In Denmark, considered a European leader in sequencing SARS-CoV-2 VoC, where testing of all positive PCR tests is commonplace, cases of Omicron have been reported to double every second day.^16^ There, almost 75% of those infected by Omicron had received full (two doses of) COVID-19 vaccination already. On the positive side, it appears most Omicron-related COVID-19 cases are mild or even pauci-symptomatic.

All viruses change in time and space by natural or artificial Darwin’s selection, and survival of the fittest,^17^ due either to high levels of herd immunity or low vaccination coverage, respectively. The toll associated with VoC Omicron underlines WHO’s COVID-19 message that “No one will be safe, until the entire World is safe (ergo vaccinated)”.

## Supporting information

TRIPOD

## Data Availability

All data produced in the present study are available upon reasonable request to the authors. We followed EQUATOR TRIPOD guidance for multivariable prediction models

https://sacoronavirus.co.za/

